# Pulmonary barotrauma in COVID-19 patients: Experience from a tertiary university hospital

**DOI:** 10.1101/2024.10.31.24316477

**Authors:** Jian Hai Chai, Azlina Masdar, Aliza Mohamad Yusof, Wan Rahiza Wan Mat

## Abstract

**Introduction**: Mechanical ventilation (MV) in COVID-19 patients is often complicated with pulmonary barotrauma. The aim of this study was to determine the incidence and risk factors associated with the development of pulmonary barotrauma in mechanically ventilated COVID-19 patients. **Materials and methods:** We included all mechanically ventilated COVID-19 patients who were aged 18 and above whom admitted to COVID ICU between January 2021 and June 2022. Patients who developed pulmonary barotrauma prior to or within 24 hours of ICU admission, iatrogenic pneumothorax, readmitted to ICU and ventilated for other causes than respiratory failure due to COVID-19 were excluded. The data for demographic, vaccination status, ventilator settings, laboratory data, steroid and immunomodulator therapies received were analysed. Univariate and multivariate analysis were carried out to determine the risk factors and outcome. **Results:** Medical records of 204 patients were included. The incidence of pulmonary barotrauma was 22.5%. Lower C-reactive protein (CRP) value on admission to ICU and FiO_2_ prescription in the first week of MV, utilisation of higher PEEP in the second week of MV and longer ventilator days predisposed patients to pulmonary barotrauma (p = 0.039, p = 0.049, p = 0.021, p = 0.036 respectively). Pulmonary barotrauma increased the duration of ICU stay (p = 0.006) and all-cause ICU mortality (p = 0.009). **Conclusion:** Lower level of CRP and FiO_2_ prescription, higher use of PEEP and longer ventilator days were the independent risk factors in our study population to develop pulmonary barotrauma which increased their duration of ICU stay and all-cause ICU mortality.

## Introduction

COVID-19 infection is caused by highly contagious SARS-CoV-2 virus and resulted in worldwide pandemic from March 2020 [1,2]. In the early phase of the infection, SARS-CoV-2 primarily targets cells, such as nasal and bronchial epithelial cells and pneumocytes, through the viral structural spike protein that binds to the angiotensin-converting enzyme 2 (ACE2) receptor. This binding facilitates viral uptake, which further promoted by type 2 transmembrane serine protease (TMPRSS2) through the cleavage of ACE2 and activating the SARS-CoV-2 S protein. Replication of viral subsequently accelerates in late stages, compromising epithelial-endothelial barrier integrity, infects pulmonary capillary endothelial cells, accentuating the inflammatory response and triggering an influx of monocytes and neutrophils, leading to the development of interstitial mononuclear inflammatory infiltrates and oedema that present as ground-glass opacities on computed tomographic (CT) imaging [3]. Liu *et al* showed that 14.1% of COVID-19 infected individuals were susceptible to developing severe disease [4]. A retrospective cohort study involving 210 patients reported that 41.8% of COVID-19 patients with severe infection progressed into acute respiratory distress syndrome (ARDS), necessitating mechanical ventilation [5]. Nevertheless, Xu *et al* reported that higher rates of ARDS were among COVID-19 patients with comorbidities [6].

Extrinsic positive end-expiratory pressure (PEEP) is often employed in mechanically ventilated patients with ARDS, to minimise lung atelectasis and thereby improve oxygenation [7]. Briel *et al* demonstrated improved rate of survival among ARDS patients when higher levels of PEEP were applied, however, with the expense of increased pulmonary barotrauma risk [8]. Pulmonary barotrauma in mechanical ventilation refers to alveolar rupture due to unrelieved pressure differential between an unvented body cavity and the surrounding air-fluid interface or across a tissue plane, leading to air leaks into extra-alveolar tissue. Pneumothorax, being the most common clinical manifestation of pulmonary barotrauma, may also present as pneumomediastinum with or without pneumothorax, as well with soft tissue extension, as subcutaneous emphysema. In COVID-19 patients requiring invasive mechanical ventilation, studies revealed that incidence of pulmonary barotrauma ranges from 9% to 33% [9–16]. Nonetheless, some authors have noted that pneumothorax or pneumomediastinum can be present independently of pulmonary barotrauma [17, 18].

Studies had shown that the incidences of pulmonary barotrauma are higher in COVID-19 infected patients, compared to non-COVID conditions, with an associated increase in mortality rate [11, 19]. Risk factors that were detected to associate with these increased incidences were among younger aged patients that had elevated lactate dehydrogenase (LDH), and mechanically ventilated using peak inspiratory pressure greater than 40 – 50 cmH_2_O, higher PEEP, higher tidal volume and higher minute ventilation, and prolonged hospital stays[9, 20–24]. Increased risk of pulmonary barotrauma was also associated with tracheal tear secondary to traumatic intubation, especially during emergency intubation, and the use of high endotracheal cuff pressure [25, 26]. Elevated endotracheal cuff pressure has been observed among COVID-19 infected patients with severe ARDS that required mechanical ventilation in the prone position [27].

While studies have yet to establish a significant correlation between comorbidities, especially pre-existing lung pathologies, with pulmonary barotrauma [28–31], the increased occurrence of pulmonary barotrauma in COVID-19 mechanically ventilated patients has been noted in our centre as well. As such, we have undertaken a study to investigate the incidence, risk factors and outcomes of these patients.

## Materials and methods

This retrospective cohort study was approved by the Research Committee of Department of Anaesthesiology & Intensive Care, Universiti Kebangsaan Malaysia Medical Centre (UKMMC) and was conducted after obtaining the approval from the Ethics Research Committee, UKM (JEP-2022-120).

Medical records of all patients that were mechanically ventilated due to COVID-19 pneumonia who were 18 years old and above, admitted to COVID intensive care unit (ICU) from January 2021 through June 2022 were recruited into this study. The COVID-19 infection was confirmed via reverse transcriptase-polymerase chain reaction (RT-PCR) and/or antigen rapid test kit (RTK-Ag), along with confirmed radiological imaging changes. Medical records of patients who developed pulmonary barotrauma prior to or within 24 hours of COVID ICU admission, had iatrogenic pneumothorax, readmission to the COVID ICU or were mechanically ventilated for reasons other than respiratory failure were excluded from the study.

Medical records of the recruited patients were retrieved from our Health Information Department, the Caring Hospital Enterprise System (C-HEts), and the Picture Archiving and Communication System (PACS) Medweb radiology portal. Data collection extended from the time of COVID ICU admission until discharge, death, or a maximum of 28 days of COVID ICU stay, whichever occurred earlier. Data that were collected were grouped into patients that developed pulmonary barotrauma and patients that did not develop this complication. The demographic profile of patients, comorbid conditions and their status of vaccination were recorded. Upon ICU admission, additional data collected were the Acute Physiology and Chronic Health Evaluation (APACHE) score, stages of ARDS following the Berlin definition of ARDS and initial laboratory investigations. ARDS severity was categorized as mild when PaO_2_/FiO_2_ ratio lies between > 200 mmHg to ≤ 300mmHg, moderate when PaO_2_/FiO_2_ ratio lies between > 100 mmHg to ≤ 200mmHg and severe when PaO_2_/FiO_2_ ratio is less than ≤ 100mmHg, with PEEP or continuous positive airway pressure (CPAP) ≥ 5cmH_2_O. Prescription of steroid and/or immunomodulator treatment, any occurrences of pulmonary embolism or pulmonary barotrauma throughout their ICU stay were collected by searching through their medical records. Confirmation of pulmonary barotrauma was done by viewing for evidence from chest X-ray (CXR), thoracic high-resolution computed tomography (HRCT) imaging or bedside lung ultrasound formal reports and/or case note documentation.

All patients recruited for this study were managed according to the lung protective strategies outlined in ARDS.net guideline to prevent ventilator-induced lung injury. We considered pulmonary barotrauma had occurred in a mechanically ventilated COVID-19 patient when pneumothorax, pneumomediastinum and/or subcutaneous emphysema was/were present. Confirmation of pulmonary barotrauma was based on imaging reports of the CXR and HRCT thorax, by the radiologist. Presence of visible thin, white line (pleural line) with absent of lung markings beyond the pleural line, rim of gas around edges of the lung, anterior of pericardium, ring around artery sign, tubular artery sign, continuous diaphragm sign and subcutaneous emphysema were among the features of arriving to the diagnosis of pulmonary barotrauma radiologically. Meanwhile, presence of subcutaneous emphysema was confirmed clinically palpating the affected area for crepitation under the skin. Bedside lung ultrasound findings, including presence of lung point, absence of lung sliding sign (or presence of barcode sign), were used by ICU consultants to support the diagnosis for patients suspected of pulmonary barotrauma. Types of pulmonary barotrauma were documented and the first incidence recorded as onset of pulmonary barotrauma. In addition, the presence of organizing pneumonia, endotracheal cuff pressure and employment of prone ventilation were also recorded.

Daily data of highest peak airway pressure (Ppeak) of the day, from day 1 to day 28 of COVID ICU admission (or day of COVID ICU discharge or death) were collected, along with the corresponding fraction of inspired oxygen concentration (FiO_2_), inspiratory pressure (IP), positive end expiratory pressure (PEEP), pressure support (PS), rate of ventilation, minute ventilation (MV) and tidal volume (TV). These data were further categorised into day 1-7, 8-14, 15 and above of COVID ICU stay. All data collected were analysed on their association in developing pulmonary barotrauma. The duration of COVID ICU stay and all-cause ICU mortality rate were recorded as outcomes associated with pulmonary barotrauma in this study. Missing or incomplete data were treated as dropouts in the analysis.

One hundred seventy medical records were the minimum sample size to be recruited with 80% power of study, a 95% confidence level and an estimated 20% dropout rate, using Krejcie & Morgan formula for finite population where number of COVID-19 cases were estimated to be of 200 [32]. Data analysis was performed using SPSS for Windows version 26.0 (IBM Corp, Armonk, NY, USA). Descriptive statistics were presented as mean ± standard deviation, median (interquartile range) or frequency (percentages) as appropriate. For between-group comparisons, independent T-test or Mann-Whitney U test was used for normally distributed continuous data and non-normally distributed data, respectively. The qualitative data was analysed using the Chi square test or Fisher Exact test, whichever appropriate. Further logistic regression (LR) analysis was performed to explore associations between pulmonary barotrauma and potential risk factors, level of inflammatory biomarkers and treatment received. Variables with p-value < 0.200 in the univariate analysis were included into the variable selection process using the backward LR method. The model fit of the final model were tested using Hosmer-Lemenshow goodness of fit test, classification tables and area under ROC curve. A p value < 0.05 was considered statistically significant.

## Results

A total of 389 patients with confirmed COVID-19 infection were admitted to our COVID ICU during the study period. Among these, 75 patients were not mechanically ventilated and 79 were admitted for reasons other than respiratory failure despite having concurrent COVID-19 infection. Additionally, 4 patients were below 18 of age, and 10 patients developed barotrauma prior to ICU admission, rendering them excluded from the study. There were 16 medical records contained incomplete data, and one patient opted for discharge at own risk from the COVID ICU, thus, these medical records were dropped out from final analysis. As a result, 204 patients’ medical record were included for data analysis.

The incidence of pulmonary barotrauma among the mechanically ventilated patients was 22.5%. Types of pulmonary barotrauma that observed were subcutaneous emphysema being the most prevalent 73.9%, followed by pneumomediastinum 65.2%, pneumothorax 52.2% and pneumopericardium 8.7%. Data on demographic, APACHE score, comorbidities, ARDS grade and status of vaccination are shown in Table 1. Most of these patients had pre-existing comorbidities, with hypertension and diabetes mellitus being the most common. Interestingly, patients who developed pulmonary barotrauma during their ICU stay were found to be significantly less critically ill upon ICU admission and had a higher prevalence of hypertension compared to those did not experience pulmonary barotrauma.

**Table 1:**
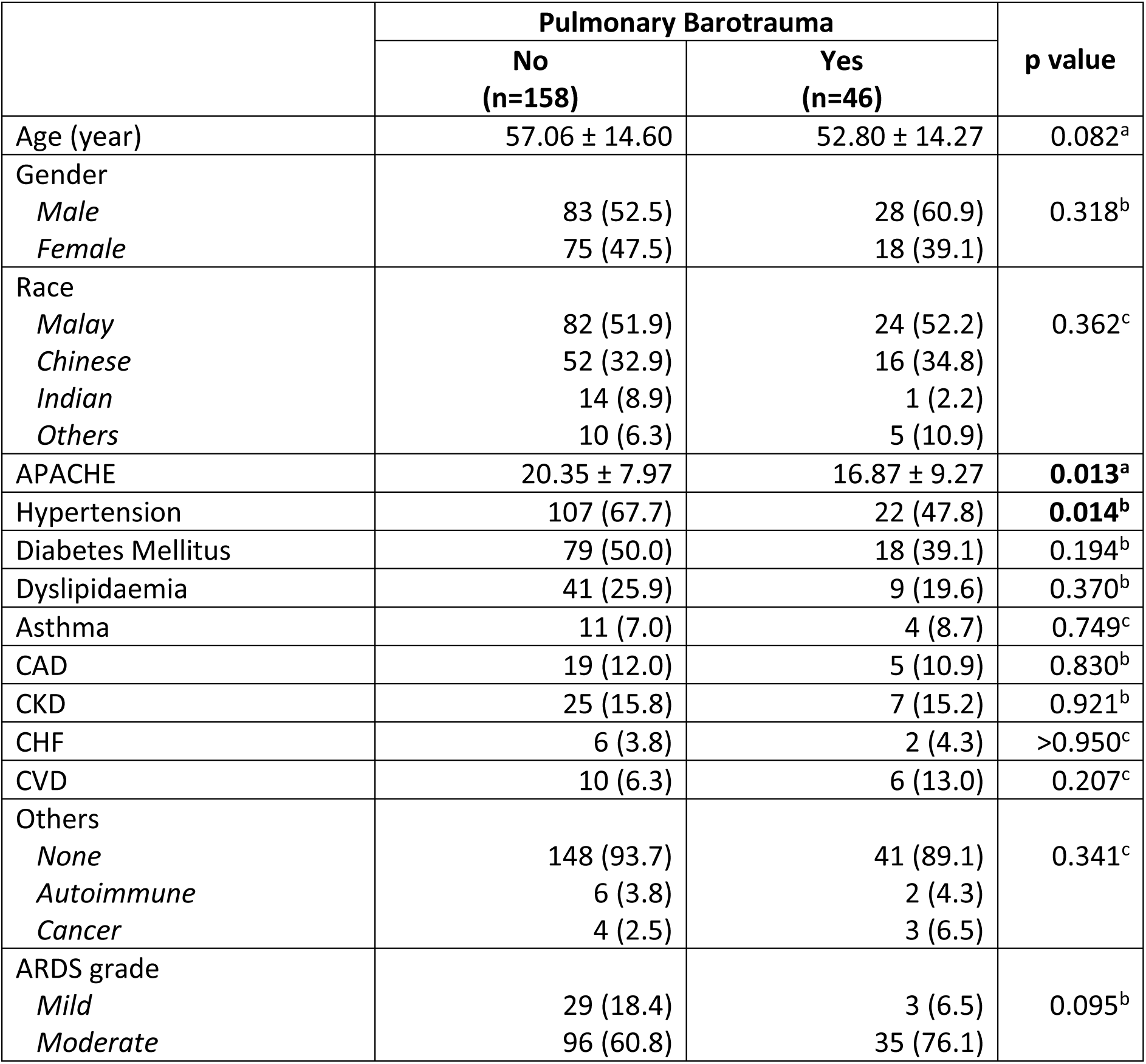
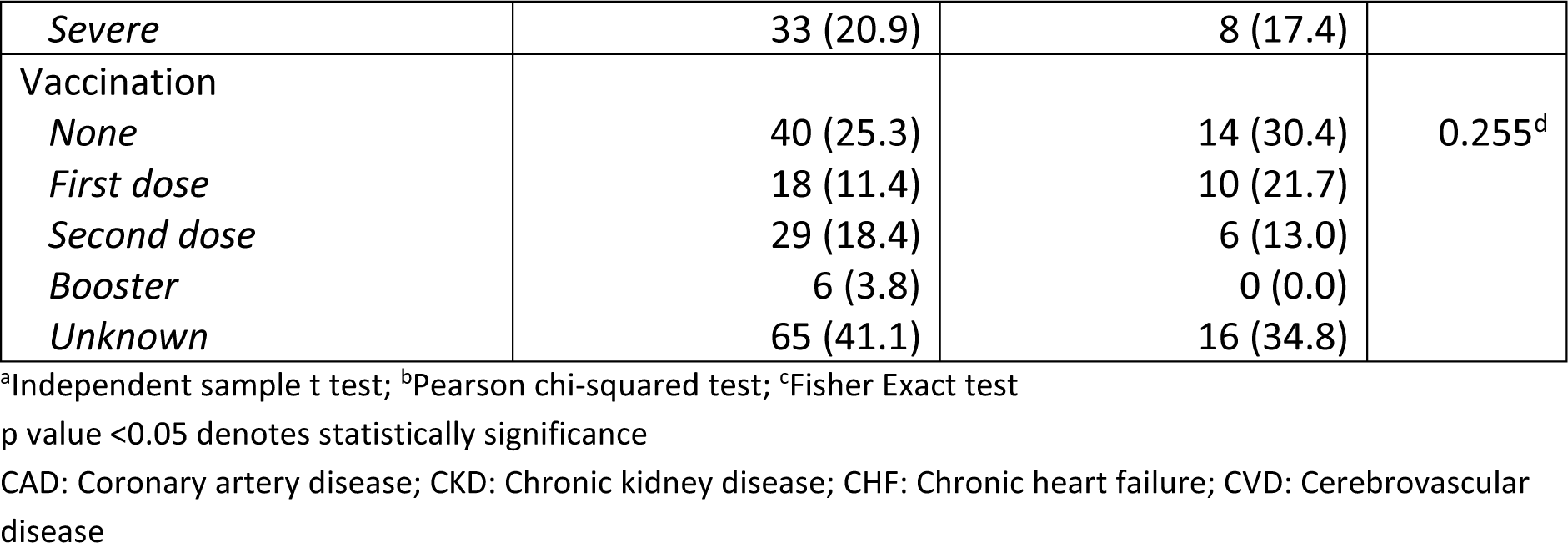
Demographic, APACHE score, comorbidities, ARDS grade and vaccination status. Data is expressed as mean ± standard deviation and number (%)

|  | Pulmonary Barotrauma |  | p value |
| --- | --- | --- | --- |
|  | No<br>(n=158) | Yes<br>(n=46) |  |
| Age (year) | 57.06 ± 14.60 | 52.80 ± 14.27 | 0.082<sup>a</sup> |
| Gender |  |  |  |
| *Male* | 83 (52.5) | 28 (60.9) | 0.318<sup>b</sup> |
| *Female* | 75 (47.5) | 18 (39.1) |  |
| Race |  |  |  |
| *Malay* | 82 (51.9) | 24 (52.2) | 0.362<sup>c</sup> |
| *Chinese* | 52 (32.9) | 16 (34.8) |  |
| *Indian* | 14 (8.9) | 1 (2.2) |  |
| *Others* | 10 (6.3) | 5 (10.9) |  |
| APACHE | 20.35 ± 7.97 | 16.87 ± 9.27 | **0.013<sup>a</sup>** |
| Hypertension | 107 (67.7) | 22 (47.8) | **0.014<sup>b</sup>** |
| Diabetes Mellitus | 79 (50.0) | 18 (39.1) | 0.194<sup>b</sup> |
| Dyslipidaemia | 41 (25.9) | 9 (19.6) | 0.370<sup>b</sup> |
| Asthma | 11 (7.0) | 4 (8.7) | 0.749<sup>c</sup> |
| CAD | 19 (12.0) | 5 (10.9) | 0.830<sup>b</sup> |
| CKD | 25 (15.8) | 7 (15.2) | 0.921<sup>b</sup> |
| CHF | 6 (3.8) | 2 (4.3) | >0.950<sup>c</sup> |
| CVD | 10 (6.3) | 6 (13.0) | 0.207<sup>c</sup> |
| Others |  |  |  |
| *None* | 148 (93.7) | 41 (89.1) | 0.341<sup>c</sup> |
| *Autoimmune* | 6 (3.8) | 2 (4.3) |  |
| *Cancer* | 4 (2.5) | 3 (6.5) |  |
| ARDS grade |  |  |  |
| *Mild* | 29 (18.4) | 3 (6.5) | 0.095<sup>b</sup> |
| *Moderate* | 96 (60.8) | 35 (76.1) |  |
| *Severe* | 33 (20.9) | 8 (17.4) |  |
| Vaccination |  |  |  |
| *None* | 40 (25.3) | 14 (30.4) | 0.255<sup>d</sup> |
| *First dose* | 18 (11.4) | 10 (21.7) |  |
| *Second dose* | 29 (18.4) | 6 (13.0) |  |
| *Booster* | 6 (3.8) | 0 (0.0) |  |
| *Unknown* | 65 (41.1) | 16 (34.8) |  |

<sup>a</sup>Independent sample t test; <sup>b</sup>Pearson chi-squared test; <sup>c</sup>Fisher Exact test
p value <0.05 denotes statistically significance
CAD: Coronary artery disease; CKD: Chronic kidney disease; CHF: Chronic heart failure; CVD: Cerebrovascular disease

**Table 2:**
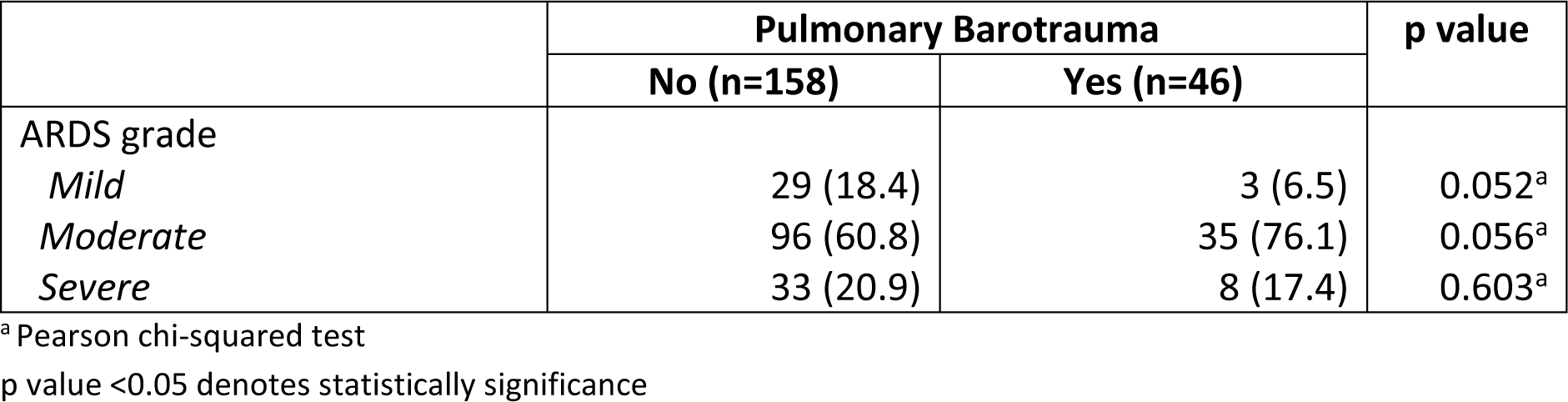
Association between ARDS grade with pulmonary barotrauma. Data is expressed as number (%)

Patients that developed pulmonary barotrauma had significantly higher PEEP applied in their second week of mechanical ventilation and had longer ventilator days as shown in Table 3.

**Table 3:**
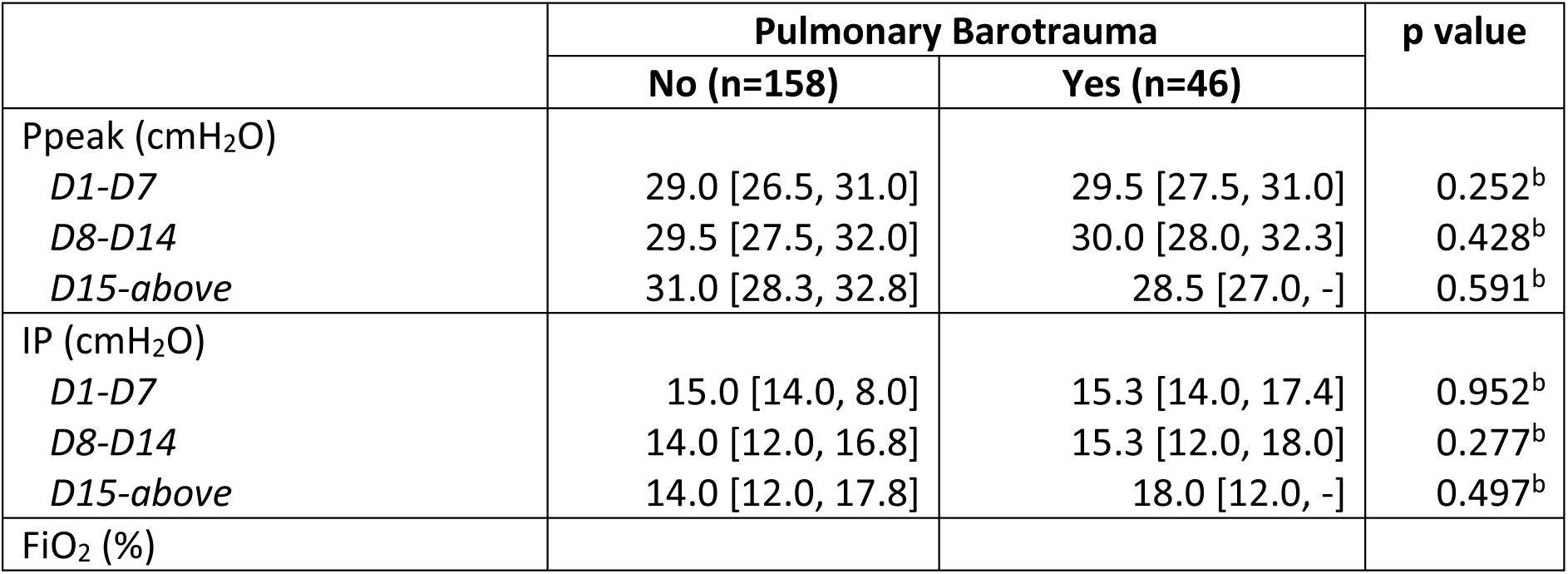
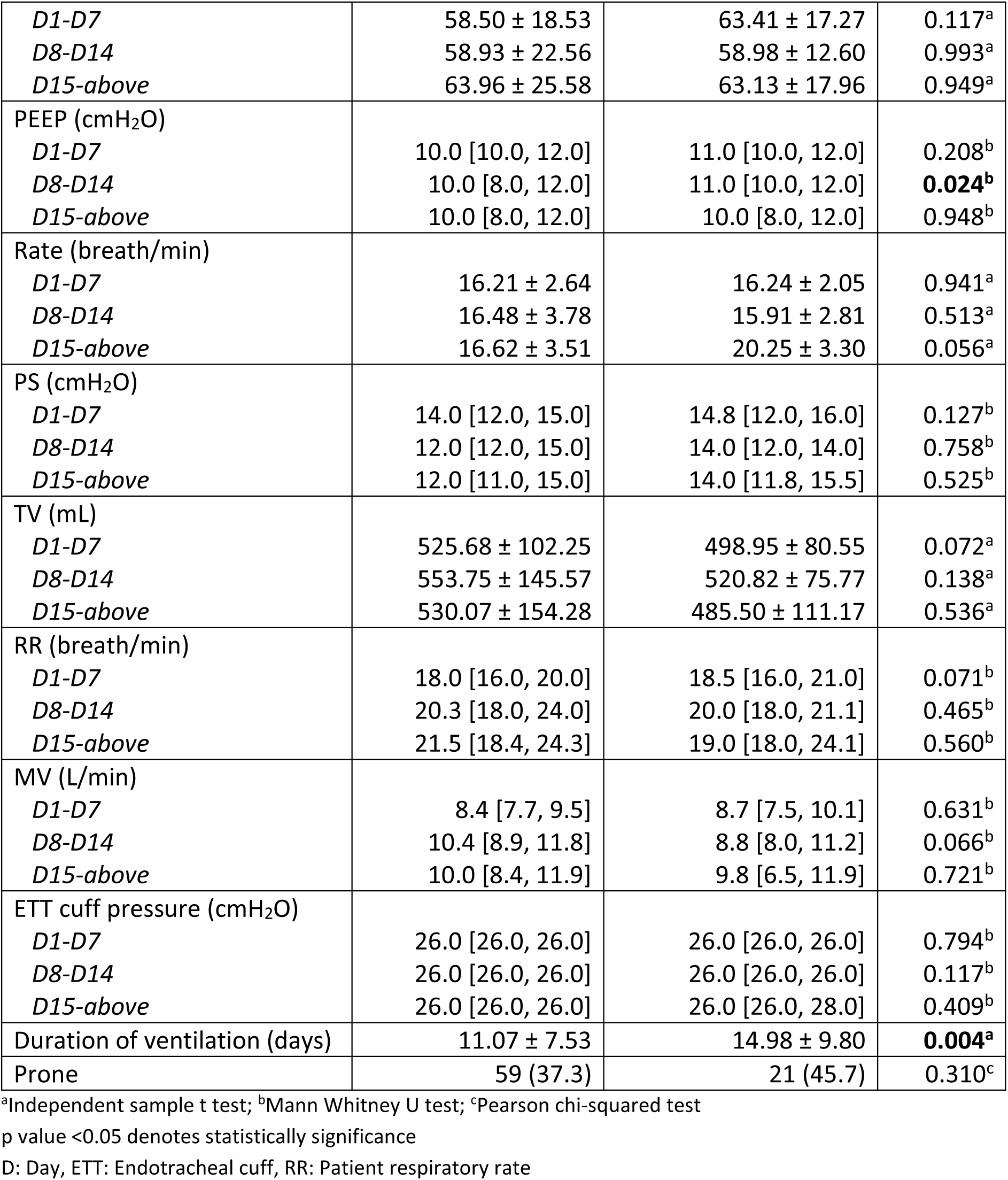
Ventilator parameters, ETT cuff pressure, prone ventilation and duration of ventilation. Data is expressed as mean ± standard deviation, median [IQR] and number (%)

|  | Pulmonary Barotrauma |  | p value |
| --- | --- | --- | --- |
|  | No (n=158) | Yes (n=46) |  |
| Ppeak (cmH<sub>2</sub>O) |  |  |  |
| *D1-D7* | 29.0 [26.5, 31.0] | 29.5 [27.5, 31.0] | 0.252<sup>b</sup> |
| *D8-D14* | 29.5 [27.5, 32.0] | 30.0 [28.0, 32.3] | 0.428<sup>b</sup> |
| *D15-above* | 31.0 [28.3, 32.8] | 28.5 [27.0, -] | 0.591<sup>b</sup> |
| IP (cmH<sub>2</sub>O) |  |  |  |
| *D1-D7* | 15.0 [14.0, 8.0] | 15.3 [14.0, 17.4] | 0.952<sup>b</sup> |
| *D8-D14* | 14.0 [12.0, 16.8] | 15.3 [12.0, 18.0] | 0.277<sup>b</sup> |
| *D15-above* | 14.0 [12.0, 17.8] | 18.0 [12.0, -] | 0.497<sup>b</sup> |
| FiO<sub>2</sub> (%) |  |  |  |
| *D1-D7* | 58.50 ± 18.53 | 63.41 ± 17.27 | 0.117<sup>a</sup> |
| *D8-D14* | 58.93 ± 22.56 | 58.98 ± 12.60 | 0.993<sup>a</sup> |
| *D15-above* | 63.96 ± 25.58 | 63.13 ± 17.96 | 0.949<sup>a</sup> |
| PEEP (cmH<sub>2</sub>O) |  |  |  |
| *D1-D7* | 10.0 [10.0, 12.0] | 11.0 [10.0, 12.0] | 0.208<sup>b</sup> |
| *D8-D14* | 10.0 [8.0, 12.0] | 11.0 [10.0, 12.0] | **0.024<sup>b</sup>** |
| *D15-above* | 10.0 [8.0, 12.0] | 10.0 [8.0, 12.0] | 0.948<sup>b</sup> |
| Rate (breath/min) |  |  |  |
| *D1-D7* | 16.21 ± 2.64 | 16.24 ± 2.05 | 0.941<sup>a</sup> |
| *D8-D14* | 16.48 ± 3.78 | 15.91 ± 2.81 | 0.513<sup>a</sup> |
| *D15-above* | 16.62 ± 3.51 | 20.25 ± 3.30 | 0.056<sup>a</sup> |
| PS (cmH<sub>2</sub>O) |  |  |  |
| *D1-D7* | 14.0 [12.0, 15.0] | 14.8 [12.0, 16.0] | 0.127<sup>b</sup> |
| *D8-D14* | 12.0 [12.0, 15.0] | 14.0 [12.0, 14.0] | 0.758<sup>b</sup> |
| *D15-above* | 12.0 [11.0, 15.0] | 14.0 [11.8, 15.5] | 0.525<sup>b</sup> |
| TV (mL) |  |  |  |
| *D1-D7* | 525.68 ± 102.25 | 498.95 ± 80.55 | 0.072<sup>a</sup> |
| *D8-D14* | 553.75 ± 145.57 | 520.82 ± 75.77 | 0.138<sup>a</sup> |
| *D15-above* | 530.07 ± 154.28 | 485.50 ± 111.17 | 0.536<sup>a</sup> |
| RR (breath/min) |  |  |  |
| *D1-D7* | 18.0 [16.0, 20.0] | 18.5 [16.0, 21.0] | 0.071<sup>b</sup> |
| *D8-D14* | 20.3 [18.0, 24.0] | 20.0 [18.0, 21.1] | 0.465<sup>b</sup> |
| *D15-above* | 21.5 [18.4, 24.3] | 19.0 [18.0, 24.1] | 0.560<sup>b</sup> |
| MV (L/min) |  |  |  |
| *D1-D7* | 8.4 [7.7, 9.5] | 8.7 [7.5, 10.1] | 0.631<sup>b</sup> |
| *D8-D14* | 10.4 [8.9, 11.8] | 8.8 [8.0, 11.2] | 0.066<sup>b</sup> |
| *D15-above* | 10.0 [8.4, 11.9] | 9.8 [6.5, 11.9] | 0.721<sup>b</sup> |
| ETT cuff pressure (cmH<sub>2</sub>O) |  |  |  |
| *D1-D7* | 26.0 [26.0, 26.0] | 26.0 [26.0, 26.0] | 0.794<sup>b</sup> |
| *D8-D14* | 26.0 [26.0, 26.0] | 26.0 [26.0, 26.0] | 0.117<sup>b</sup> |
| *D15-above* | 26.0 [26.0, 26.0] | 26.0 [26.0, 28.0] | 0.409<sup>b</sup> |
| Duration of ventilation (days) | 11.07 ± 7.53 | 14.98 ± 9.80 | **0.004<sup>a</sup>** |
| Prone | 59 (37.3) | 21 (45.7) | 0.310<sup>c</sup> |

<sup>a</sup>Independent sample t test; <sup>b</sup>Mann Whitney U test; <sup>c</sup>Pearson chi-squared test
p value <0.05 denotes statistically significance
D: Day, ETT: Endotracheal cuff, RR: Patient respiratory rate

Laboratory data collected on the day of ICU admission, along with presence of organizing pneumonia (OP), pulmonary embolism (PE) and the administration of steroid and immunomodulator are presented in Table 4. Interestingly, patients with significantly elevated CRP levels and those diagnosed with PE were less likely to develop pulmonary barotrauma.

**Table 4:** Biochemical, lung pathology, the use of steroid and immunomodulator. Data is expressed as median [IQR] and number (%)

|  | Pulmonary Barotrauma |  | p value |
| --- | --- | --- | --- |
|  | No<br>(n=158) | Yes<br>(n=46) |  |
| Lymphocyte (X 10<sup>9</sup>/L) | 0.80 [0.50, 1.03] | 0.60 [0.50, 1.10] | 0.418<sup>a</sup> |
| Platelet (X 10<sup>9</sup>/L) | 237.0 [194.5, 303.0] | 266.0 [191.0, 327.3] | 0.341<sup>a</sup> |
| CRP (mg/dL) | 8.53 [3.84, 17.20] | 5.87 [2.76, 11.45] | **0.033<sup>a</sup>** |
| Ferritin (mcg/L) | 1424.15<br>[767.62, 3049.85] | 2035.16<br>[1024.58, 3068.86] | 0.222<sup>a</sup> |
| D-dimer (g/L) | 2.11 [1.05, 3.99] | 2.18 [0.82, 5.18] | 0.788<sup>a</sup> |
| LDH (IU/L) | 562.50<br>[440.75, 764.00] | 680.00<br>[487.00, 890.00] | 0.078<sup>a</sup> |
| OP | 97 (61.4) | 32 (69.6) | 0.312<sup>b</sup> |
| PE | 47 (29.7) | 6 (13.0) | **0.023<sup>b</sup>** |
| Use of steroid | 156 (98.7) | 46 (100.0) | >0.950<sup>c</sup> |
| Use of immunomodulator | 114 (72.2) | 29 (63.0) | 0.235<sup>b</sup> |

<sup>a</sup>Mann Whitney U test; <sup>b</sup>Pearson chi-squared test; <sup>c</sup>Fisher Exact test
p value <0.05 denotes statistically significance
CRP: C-reactive protein

Logistic regression analysis was performed to explore the factors associated with pulmonary barotrauma. Our univariate analysis revealed that a higher APACHE score upon COVID ICU admission, the presence of hypertension and PE were associated with lesser risk of pulmonary barotrauma. In contrast, the presence of ARDS, increased PEEP use during the second week of MV and prolonged ventilation duration posted significant greater risk of pulmonary barotrauma as detailed in Table 5. Further insights from multivariate analysis indicated that patients with lower CRP levels upon ICU admission, higher FiO_2_ requirements during the first week of MV, increased PEEP use during the second week of MV and longer ventilated days were observed to increase their odds for developing pulmonary barotrauma.

**Table 5:** Risk factors for pulmonary barotrauma.

|  | Univariable analysis |  | Multivariable analysis |  |
| --- | --- | --- | --- | --- |
|  | OR (95% CI) | p value | Adj. OR (95% CI) | p value |
| Age | 0.98 [0.96, 1.00] | 0.084 |  |  |
| Gender |  |  |  |  |
| Male | Ref |  |  |  |
| Female | 0.71 [0.36, 1.39] | 0.319 |  |  |
| Race |  |  |  |  |
| Malay | Ref |  |  |  |
| Chinese | 1.05 [0.51, 2.16] | 0.892 |  |  |
| Indian | 0.24 [0.03, 1.95] | 0.184 |  |  |
| Others | 1.71 [0.53, 5.48] | 0.368 |  |  |
| APACHE | 0.95 [0.91, 0.99] | 0.014 |  |  |
| Hypertension | 0.44 [0.22, 0.85] | 0.015 |  |  |
| Diabetes | 0.64 [0.33, 1.26] | 0.196 |  |  |
| Dyslipidaemia | 0.69 [0.31, 1.56] | 0.377 |  |  |
| Asthma | 1.27 [0.39, 4.20] | 0.692 |  |  |
| CAD | 0.89 [0.31, 2.54] | 0.831 |  |  |
| CKD | 0.96 [0.38, 2.37] | 0.921 |  |  |
| CHF | 1.15 [0.22, 5.91] | 0.866 |  |  |
| CVD | 2.22 [0.76, 6.48] | 0.144 |  |  |
| Others |  |  |  |  |
| None | Ref |  |  |  |
| Autoimmune | 1.20 [0.23, 6.19] | 0.825 |  |  |
| Cancer | 2.71 [0.58, 12.58] | 0.204 |  |  |
| ARDS grade |  |  |  |  |
| Mild | Ref |  |  |  |
| Moderate | 3.52 [1.01, 12.30] | 0.048 |  |  |
| Severe | 2.34 [0.57, 9.67] | 0.239 |  |  |
| Vaccination |  |  |  |  |
| None | Ref |  |  |  |
| First dose | 1.59 [0.59, 4.25] | 0.357 |  |  |
| Second dose | 0.59 [0.20, 1.72] | 0.335 |  |  |
| Booster | - | - |  |  |
| Unknown | 0.70 [0.31, 1.59] | 0.399 |  |  |
| Lymphocyte | 0.89 [0.53, 1.49] | 0.653 |  |  |
| Platelet | 1.00 [1.00, 1.01] | 0.262 |  |  |
| CRP | 0.97 [0.93, 1.01] | 0.153 | 0.90 [0.81, 0.99] | **0.039** |
| Ferritin | 1.00 [1.00, 1.00] | 0.801 |  |  |
| D-dimer | 1.00 [0.99, 1.01] | 0.707 |  |  |
| LDH | 1.00 [1.00, 1.00] | 0.850 |  |  |
| OP | 1.44 [0.71, 2.91] | 0.313 |  |  |
| PE | 0.35 [0.14, 0.89] | 0.028 | 0.33 [0.08, 1.31] | 0.114 |
| Prone | 1.41 [0.73, 2.74] | 0.311 |  |  |
| Ppeak |  |  |  |  |
| D1-D7 | 1.04 [0.96, 1.13] | 0.341 |  |  |
| D8-D14 | 1.05 [0.91, 1.22] | 0.485 |  |  |
| D15-above | 0.94 [0.70, 1.27] | 0.699 |  |  |
| IP |  |  |  |  |
| D1-D7 | 0.99 [0.89, 1.09] | 0.831 |  |  |
| *D8-D14* | 1.02 [0.91, 1.15] | 0.751 |  |  |
| *D15-above* | 1.08 [0.85, 1.37] | 0.534 |  |  |
| FiO<sub>2</sub> |  |  |  |  |
| *D1-D7* | 1.01 [1.00, 1.03] | 0.119 | 0.95 [0.90, 0.99] | **0.049** |
| *D8-D14* | 1.00 [0.98, 1.02] | 0.993 |  |  |
| *D15-above* | 1.00 [0.96, 1.04] | 0.948 |  |  |
| PEEP |  |  |  |  |
| *D1-D7* | 1.10 [0.91, 1.34] | 0.310 |  |  |
| *D8-D14* | 1.37 [1.02, 1.84] | 0.039 | 1.68 [1.08, 2.61] | **0.021** |
| *D15-above* | 1.00 [0.63, 1.59] | >0.950 |  |  |
| Rate |  |  |  |  |
| *D1-D7* | 1.01 [0.88, 1.15] | 0.940 |  |  |
| *D8-D14* | 0.95 [0.83, 1.10] | 0.510 |  |  |
| *D15-above* | 1.35 [0.97, 1.87] | 0.072 |  |  |
| PS |  |  |  |  |
| *D1-D7* | 1.09 [0.96, 1.25] | 0.192 |  |  |
| *D8-D14* | 1.04 [0.86, 1.26] | 0.664 |  |  |
| *D15-above* | 1.18 [0.76, 1.84] | 0.458 |  |  |
| TV |  |  |  |  |
| *D1-D7* | 1.00 [0.99, 1.00] | 0.113 |  |  |
| *D8-D14* | 1.00 [0.99, 1.00] | 0.304 |  |  |
| *D15-above* | 1.00 [0.99, 1.00] | 0.528 |  |  |
| RR |  |  |  |  |
| *D1-D7* | 1.09 [0.98, 1.21] | 0.115 |  |  |
| *D8-D14* | 0.95 [0.84, 1.07] | 0.398 |  |  |
| *D15-above* | 0.94 [0.73, 1.20] | 0.599 |  |  |
| MV |  |  |  |  |
| *D1-D7* | 1.04 [0.85, 1.26] | 0.724 |  |  |
| *D8-D14* | 0.83 [0.67, 1.04] | 0.106 |  |  |
| *D15-above* | 0.92 [0.64, 1.31] | 0.640 |  |  |
| ETT |  |  |  |  |
| *D1-D7* | 0.73 [0.21, 2.57] | 0.626 |  |  |
| *D8-D14* | 0.27 [0.03, 2.62] | 0.261 |  |  |
| *D15-above* | 2.02x10<sup>14</sup> [0.00, ∞] | 0.998 |  |  |
| Duration of ventilation | 1.05 [1.02, 1.10] | 0.006 | 1.08 [1.01, 1.16] | **0.036** |
| Use of steroid | - | 0.999 |  |  |
| Use of immunomodulator | 0.66 [0.33, 1.32] | 0.237 |  |  |

p value <0.05 denotes statistical significance
Variables with p-value < 0.200 in the univariate analysis were included into the variable selection process using backward LR method.

Patients who developed pulmonary barotrauma had a significantly longer stayed in COVID ICU by approximately 5 days (mean duration of ICU stays in patients with pulmonary barotrauma 17.07±10.53 days vs patients that did not develop pulmonary barotrauma 12.34±7.26 days, p = 0.006). Furthermore, patients with pulmonary barotrauma were associated with a markedly higher all-cause ICU mortality rate compared to those without pulmonary barotrauma (67.4% *vs* 32.6%, p = 0.009).

## Discussion

The limited understanding towards this novel and yet rapidly evolving COVID-19 virus, coupled with heterogenous findings from published studies, had however hindered the progression on identifying an ideal treatment. Pulmonary barotrauma has been closely linked with increased morbidity and mortality [33] due to hypoxia and hypotension secondary to obstructive shock in cases of tension pneumothorax. We found the incidence of pulmonary barotrauma was 22.5%, closely aligning with Belletti A *et al* at 24.1% [17]. Lower incidence rate, on the other hand, was reported by Edwards JA *et al* [43] (9.4%) while Kahn MR *et al* [14] observed a higher rate (33.3%). Nonetheless, other studies documented consistent range of incidence rate between 13-14.8% [10, 11, 13, 15, 20, 24, 47]. It is interesting to note that elevated rates of pulmonary barotrauma were observed during previous SARS and MERS coronavirus outbreak, ranging between 12-34%, raising questions whether viral infections, or more specifically coronavirus infections pose a unique risk [30–32, 34].

While pre-existing lung diseases are traditionally associated with the development of pulmonary barotrauma during mechanical ventilation [35], we found no significant correlation between comorbidities and pulmonary barotrauma in our COVID-19 patient cohort, consistent with other studies examining COVID-19 patients [13, 14, 17, 36, 37].

A key finding in our study was that lower CRP values on COVID ICU admission were found to pose a higher risk in developing pulmonary barotrauma. Severe COVID-19 disease is linked to a hyperinflammatory state with release of various cytokines, including IL-6, proposing the inhibition of IL-6 a potential therapeutic target. However, it is important to note that IL-6 also plays a key role in viral clearance and angiogenesis. Inhibition of IL-6 could potentially cause harm by hindering viral clearance and interrupts the remodelling and repair process that prevents pulmonary barotrauma. Taha M *et al* [37] reported increased pneumothorax rate in recipients of Tozilizumab, an immunomodulator that also prescribed in our patients. Despite that, we cannot rule out the possibility that barotrauma occurred late in the disease course, possibly after CRP levels had already declined. In addition, steroid was prescribed to almost entire population of our study subjects, with larger doses used on patients with severe hypoxemia, potentially contributed to the lower CRP value obtained in our study.

Despite implementing lung protective strategy in ventilating ARDS patients, studies on COVID-19 continue to demonstrate a higher incidence of pulmonary barotrauma among COVID-19 patients compared to non-COVID-19 ventilated patients [10, 15, 16, 38, 39]. In contrast to other studies that found no difference in ventilator settings, we observed that lower FiO_2_ prescription during the first week of mechanical ventilation and a higher PEEP value during the second week were risk factors for barotrauma [14]. It was postulated that lung pathophysiology in COVID-19 may differ from typical pneumonia. COVID-19-induced ARDS may exhibit 2 distinct phenotypes, namely “L” and “H” type. Patients with type “L” ARDS present with low elastance and low response to PEEP due to absence of recruitable alveoli, whereby those with type “H” are characterised as high elastance, low lung volume and high PEEP response, resembling “classic” ARDS. Vasoplegia in type “L” ARDS is responsible for severe hypoxaemia seen in COVID-19 patients, which responded by increasing minute ventilation, primarily by increasing the tidal volume. This led to more negative intrathoracic inspiratory pressure, resulting in patient self-inflicted lung injury (P-SILI), which predisposes them to pulmonary barotrauma [40–42]. We hypothesize that increasing FiO_2_ for type “L” patients could potentially mitigate P-SILI, thereby reducing the risk of pulmonary barotrauma.

The virus-induced damage to ACE2 receptor in endothelial and epithelial cells may impair the cellular repair mechanism of the lung, exacerbating alveolar injury [43, 44]. Nevertheless, the interplay between cytokine production, PEEP and tidal volume may worsen alveolar injury by amplifying the inflammatory response, predisposing patients to pulmonary barotrauma [45]. This finding suggests that a deeper understanding on state of the disease itself may contribute higher weightage in preventing pulmonary barotrauma than mechanical ventilator settings alone. Supporting this, several studies published have found no association between ventilator settings and pulmonary barotrauma incidence [14, 15, 17, 37, 38], and in fact, Lemmers DH *et al* [15] reported that PEEP and plateau pressure were lower on the day pulmonary barotrauma occurred than on the day of ICU admission. Nonetheless, heightened respiratory drive in COVID-19 patients may induce self-inflicted lung injury due to heterogenous alveolar strain, contributed to the development of pulmonary barotrauma, explaining the occurrence of incident between day 8-14 as the disease progresses [46].

Duration of ventilation also found to be linked with pulmonary barotrauma risk, which also reported by Taha M *et al* [37]. This is consistent with the reported finding of higher frequency of pulmonary barotrauma in mechanical ventilated patients than non-mechanical ventilated patients in other studies [16, 30]. Nonetheless, mechanical ventilation itself increases the risk of macro- and microbarotrauma, driven by positive pressure ventilation, recruitment manoeuvres, coughing, and ventilator asynchrony [48, 49]. Our study corroborates previous research showing that patients who develop barotrauma experience longer ICU stays and higher mortality rates. Specifically, we observed a 67.4% mortality rate among patients with barotrauma, which aligns with findings by Dubey R *et al* and Kahn MR *et al* [13, 14].

This study has limitations inherent to its retrospective design, including missing, misclassified, and non-retrievable data, as information was manually documented by healthcare personnel. Future studies could benefit from a prospective, multi-centre design to enhance the generalisability of results. Additionally, unmeasured confounding factors may have influenced our findings. As we continue to explore potential risk factors for pulmonary barotrauma, our study provides a valuable foundation for future research and guidance for clinicians seeking to optimize patient care.

## Conclusion

Pulmonary barotrauma occurred in nearly a quarter of our COVID-19 ICU patients. Independent risk factors identified in our study included lower CRP levels, lower FiO_2_ prescription in the first week of mechanical ventilation, higher PEEP prescription in the second week, and longer ventilation duration. These factors were associated with prolonged ICU stays and significantly elevated mortality rates. Our findings underscore the need for improved disease understanding and vigilant patient management to mitigate the risk of pulmonary barotrauma in COVID-19 patients.

## Data Availability

All data produced in the present work are contained in the manuscript

## Acknowledgement

I sincerely thank Associate Professor Dr. Azlina Masdar, consultant anaesthesiologist at the Department of Anaesthesiology and Intensive Care, Universiti Kebangsaan Malaysia Medical Centre (UKMMC), for her invaluable guidance and support throughout this study. I am also grateful to Associate Professor Dr. Aliza Mohamad Yusof, consultant intensivist, for her insightful advice during the writing process. My heartfelt appreciation goes to the lecturers and colleagues at UKMMC for their assistance and encouragement. Finally, I extend my deepest gratitude to my family for their unwavering love and support, which made this work possible.

